# Examining the effect of information channel on COVID-19 vaccine acceptance

**DOI:** 10.1101/2021.01.18.21250049

**Authors:** R. Piltch-Loeb, E. Savoia, B. Goldberg, B. Hughes, T. Verhey, J. Kayyem, C. Miller-Idriss, MA. Testa

## Abstract

Hesitancy towards the COVID-19 vaccine remains high among the US population. Now that the vaccine is available to priority populations, it is critical to convince those that are hesitant to take the vaccine. Public health communication about the vaccine as well as misinformation on the vaccine occurs through a variety of different information channels. Some channels of information are more commonly found to spread misinformation. Given the expansive information environment, we sought to characterize the use of different media channels for COVID-19 vaccine information and determine the relationship between information channel and vaccine acceptance. We conducted a convenience sample of vaccine priority groups (N=2,650) between December 13 and 23, 2020 and conducted bivariate chi-squared tests and multivariable multinomial logistic regression analyses to determine the relative impact of channels of information on vaccine acceptance. We found traditional channels of information, especially National TV, National newspapers, and local newspapers increased the relative risk of vaccine acceptance. Individuals who received information from traditional media compared to social media or both traditional and social media were most likely to accept the vaccine. The implications of this study suggest social media channels have a role to play in educating the hesitant to accept the vaccine, while traditional media channels should continue to promote data-driven and informed vaccine content to their viewers.

## Introduction

The COVID-19 vaccine has been met with a mix of excitement and apprehension. Vaccine hesitancy towards a COVID-19 vaccine remains high in the United States, with close to 40% of the population unsure if they will be vaccinated, and another 30% unlikely to do so as of the end of 2020 (1). The level of vaccine hesitancy in anticipation of vaccine approval by regulatory agencies has become more critical now that the vaccine is available, especially for high priority health care workers and vulnerable populations. The control of COVID-19 is largely dependent upon its uptake. Accordingly, the impact of public risk communication efforts on the effectiveness, safety and availability of the vaccine are critical to vaccine uptake. Furthermore, these efforts can be implemented through numerous information channels. How these channels are perceived by the public and which channels are used to receive information about both COVID-19 and the vaccine will determine the likelihood of achieving satisfactory levels of vaccine uptake (2).

Since the beginning of the COVID-19 pandemic, there has been an abundance of information, with nearly every media channel covering the latest developments (3). These channels are sharing vaccine information and may be very influential in swaying public opinion as to whether or not they want to be vaccinated. Past research has found that online forums, blogs, and social media have contributed to the spread of vaccine hesitancy. Several authors have examined how social media platforms contribute to vaccine hesitancy such as by promoting personal narratives over empirical data and connecting anti-vaccination themes to broader belief systems of freedom of choice and parental rights (4-7). Social media has continued to be a vehicle for the spread of COVID-19 misinformation (2).

Misinformation related generally to the pandemic at large has spread online, and has increased with the introduction of the vaccine (2). When the pandemic began, misinformation focused on various theories about the virus as discussed by Singh et al., 2020. One theory hypothesized that the virus was first transmitted from bats to humans in a Wuhan, China, wet market (8). A second theory suggested that poor quality control during virology research experiments led to the escape of the virus from China’s first biosafety level 4 (BSL–4) laboratory located in Jiangxia District, Wuhan, Hubei. A third theory attributed the spread of the virus to a bioweapons military project program being conducted by the Chinese Government. Singh and colleagues (2020) found that low quality sources that were unverified and appeared to promote misinformation on COVID-19 were more commonly retweeted than those with high quality information linked to verifiable health authorities. In a comparative analysis of the spread of COVID-19 misinformation on five social media platforms (Twitter, Instagram, YouTube, Reddit and Gab), Cinelli and colleagues (2020) analyzed more than 8 million comments and posts to model the spread of misinformation (9). They found that amplification of misinformation varied by platform, suggesting that misinformation dissemination depends on each platform’s interaction paradigms and by the specific interaction patterns of groups of users engaged with the topic.

The content and spread of misinformation online can influence risk perception and vaccine hesitancy (10, 11). Exposure to websites and social media posts discouraging immunizations, even when brief, can increase perception of vaccination risk while decreasing the benefits (10). Viewing such content can reduce viewers’ vaccination intentions (11). Buller and colleagues (2019) describe how Facebook users appeared disproportionately swayed towards vaccine hesitancy due to narratives and emotional stories(12). Nadler, Crain and Donovan (2018) describe this advantage of poor-quality information over good as a direct and predictable consequence of the digital media ecosystem in combination with practices and technologies of consumer monitoring, audience customization, and the automation of influence campaigns (13).

Individuals tend to receive information from multiple information channels during a pandemic (14, 15). Bennet, Manheim and others, suggests that patterns of homogenization, polarization, and targeted marketing have created a “one-step flow” of persuasion even in legacy media, in which interpersonal social influence recedes as targeted media increasingly exercises direct influence over individual opinion (16-18). There is much discussion and research on social media as a vehicle for misinformation and vaccine skepticism, and less on the role that traditional media, such as TV, newspapers, and radio can play in misinformation (19). An individual’s perception of a given channel can impact how they act upon that information. Perceptions of credibility, authoritativeness, and persuasiveness vary by information source (20, 21). For example, information that is seen as straightforward, not sensationalized, and accurate from legacy and local news outlets has been shown to increase vaccine acceptance (22).

Given the expansive information environment and need for vaccine uptake, we sought to characterize the use of different media channels for COVID-19 vaccine information, how that information is trusted, and the relative effect on vaccination likelihood. We specifically had two research questions, namely:

1. How much do individuals trust vaccine information from different information channels?
2. What is the relationship between channels of information and vaccine acceptance?

We hypothesized that information from traditional media would be more positive and trusted compared to social media channels, and that getting information from traditional media channels would be more likely to increase willingness to take the vaccine.

## Methods

### Study Design

We conducted a cross-sectional online survey via mobile devices on the Pollfish survey platform. Pollfish pays mobile application developers to display the surveys within their applications. To incentivize participation, small monetary incentives are provided to randomly selected users who complete the surveys. An initial survey draft underwent pilot cognitive testing with 20 individuals, and the survey was subsequently revised for length and clarity. The survey was designed to be completed in approximately ten minutes or less and for ease of completion on a mobile device. The survey was conducted between December 13^th^ and 23^rd^, 2020.

Respondents were required to be healthcare workers or in a potential vaccine priority group based on national guidance available at the time the survey was developed (23). Respondents were also required to live in the United States. The study protocol and survey instrument were approved by the Harvard T.H. Chan School of Public Health Institutional Review Board.

### Dependent Variable

Respondents were asked about their intentions to take the vaccine by answering the question how likely they would be to receive a COVID-19 vaccine if offered to them at no cost within two months. The six response options were very likely, somewhat likely, somewhat unlikely, very unlikely, would consider it after two months, and not sure. For the scope of this analysis, the six response options were categorized into the dependent variable of “vaccine acceptance”, on the rating scale defined as: 1) “vaccine acceptant”-those that reported “very likely”, 2) “some hesitancy” in taking the vaccine-those that reported they were “somewhat likely”, “somewhat unlikely”, “would get it after two months”, or were “not sure”, and 3) “vaccine hesitant”-those who were “very unlikely.”

### Independent variables

The effects of COVID-19 vaccine information channels, and the trust in information from those channels, on vaccine acceptance is the primary interest of this analysis. Respondents were asked to select up to three channels from which they had received most of their information on the COVID-19 vaccine. Since information channels are not mutually exclusive, each channel was treated as an indicator variable, dichotomized by whether or not the channel had been selected. In addition, we also created a variable to characterize the sources of information an individual selected with three distinct categories, namely: getting most information from only traditional media channels (TV, newspaper, or radio); getting most information from only social media channels (Facebook, Instagram, YouTube, Twitter, or Tiktok); and getting most information from both social and traditional media channels.

Trust in information was measured by analyzing responses to the question “*How much do you trust the information you got so far about the COVID-19 vaccine?*” with response options: not at all, very little, somewhat, and a lot. Not at all, and very little were combined into a category named as “low trust” to obtain an ordered categorical “trust” variables with three levels, low trust, some trust, and high trust. Respondents also selected each channel of COVID-19 vaccine information they had been exposed to overall and rated the quality of information received from that channel as positive, negative, or neutral.

In addition to the primary independent variables of interest described above - information channels accessed and trust in information - we also explored several independent variables that have previously been found to relate to vaccine hesitancy and health behaviors including socio-demographics, perceived risk, COVID-19 experience, and prior vaccination behaviors. Socio-demographic variables were age, gender, race, level of education, and type of job (working in the healthcare sector versus not working in this sector). Age categories were re-coded to combine small sample subgroups for age groups over 55. Lower education levels were collapsed to create one category that referenced high school education or lower and General Education Development (GED) or equivalent. COVID-19 risk perception was measured by asking respondents to report their level of concern of contracting COVID-19 at work or outside their work environment, and their level of concern about infecting family members or friends. Responses were dichotomized into two risk-related variables (medium/low reported risk vs. high risk). COVID-19 experience included having been sick with COVID-19 (yes/no), knowing someone who had a severe COVID-19 infection (yes/no), and knowing someone who had died of COVID-19 (yes/no). As a proxy for prior vaccination behavior, we asked respondents if they had received their flu vaccine this year.

### Statistical Analysis

To answer our first research question regarding the level of trust in COVID-19 information from various channels, we analyzed the proportion of respondents who utilized each information channel, the quality rating of vaccine information from each channel, as well as the level of trust per channel using descriptive statistics. In preparation for multivariable analysis, and to answer our second research question concerning acceptance of the vaccine, we conducted bivariate analyses using chi-squared tests and multivariable regression analysis to study the association between the independent variables and the likelihood of taking the COVID-19 vaccine. We conducted chi-squared tests of independence to test for associations between the levels of each independent variable and the levels of the dependent variable indicative of vaccine hesitancy, some hesitancy, or acceptant. We used a p-value of <0.05 as the threshold for inclusion of the predictor variable in the multinomial logistic regression model. We also tested for collinearity among predictors by Spearman’s rank correlation test prior to the completion of the regression analysis, using the cutoff of 0.7 as a measure of collinearity. We specifically tested for collinearity among information sources and found that the degree of collinearity was not high enough to exclude a particular source. Due to the significant challenges in convincing those who are strongly committed in their anti-vaccination beliefs (24, 25), for multivariable regression analysis we conducted multinomial logistic regression to identify the relative risk of being very likely to take the vaccine compared to those in the some-hesitancy group. A generalized Hosmer–Lemeshow goodness-of-fit test for multinomial logistic regression models was used to assess the goodness of fit (26). Data analysis was performed using Stata Statistical Software: Release 16 College Station, TX: StataCorp LLC.

## Results

### Sample characteristics

There were 2,650 in our analytic sample. The sample was predominantly White 66.0% White, male (53.9%), and 65.9% were between 25 and 44 years of age. The majority worked in the healthcare sector (61.4%) and were highly education with 54.6% possessing a Bachelor’s degree or higher. In terms of COVID-19 experience, approximately one-quarter of respondents reported having COVID-19, 22.6% had a close family member or friend who was a severe case, and 11.3% had a close family member or friend who had died of COVID-19. The prevalence of COVID in the sample was over three times that reported in the United States (23,214,472 cases out of a population of 331,992,651 as of January 12, 2021) reflecting the intended high exposure and infection rate in the sample. The majority of respondents had a high level of perceived risk of contracting COVID-19 at work or outside of work (65.3%) and infecting others with COVID-19 (62.5%). About half of the sample had received their flu vaccine. Among all respondents, 12.8% were very unlikely to take the vaccine in the next two months 47.3% had some level of hesitancy, and 39.9% were very likely to do so. A detailed breakdown of these characteristics is given in Table 2.

**TABLE 1.** Description of COVID-19 information from different channels.

Effect of information channel on COVID-19 vaccine acceptance
| TABLE 1. Description of COVID-19 information from different channels |  |  |  |  |  |  |  |
| --- | --- | --- | --- | --- | --- | --- | --- |
|  | Users<br>n (%) | Quality of information<br>n (%) |  |  | Level of trust in vaccine information from<br>channel<br>n (%) |  |  |
|  | 2,650<br>(100%) | Negative | Positive | Neutral | Low | Some | High |
| Overall |  | 429 (16.3) | 1,411<br>(53.6) | 666 (25.3) | 916 (36.3) | 1,023 (40.5) | 584 (23.1) |
| By channel |  |  |  |  |  |  |  |
| National TV | 896 (33.8) | 202 (22.7) | 418 (47.0) | 205 (23.0) | 275 (30.7) | 355 (39.6) | 266 (29.7) |
| Local TV | 1,629 (61.3) | 143 (8.8) | 986 (60.7) | 463 (28.5) | 574 (35.3) | 667 (40.9) | 388 (23.8) |
| National Newspaper | 608 (22.9) | 144 (23.7) | 293 (48.3) | 128 (21.1) | 169 (27.8) | 232 (38.2) | 207 (34.1) |
| Local Newspaper | 335 (12.6) | 75 (22.4) | 190 (56.7) | 53 (15.8) | 103 (30.7) | 134 (40.0) | 98 (29.3) |
| Radio | 356 (13.4) | 95 (26.7) | 172 (48.3) | 63 (17.7) | 100 (28.1) | 135 (37.9) | 121 (34.0) |
| Facebook | 1,006 (37.8) | 159 (15.8) | 645 (64.1) | 202 (20.1) | 383 (40.7) | 370 (39.3) | 189 (20.1) |
| YouTube | 758 (28.5) | 116 (15.3) | 519 (68.5) | 123 (16.2) | 305 (42.5) | 285 (39.7) | 127 (17.7) |
| Instagram | 574 (21.6) | 90 (15.7) | 397 (69.2) | 87 (15.2) | 233 (43.1) | 201 (37.2) | 106 (19.6) |
| Twitter | 485 (18.2) | 58 (12.0) | 362 (74.6) | 65 (13.4) | 203 (44.4) | 170 (37.2) | 84 (18.4) |
| Tiktok | 298 (11.2) | 71 (23.8) | 172 (57.7) | 55 (18.5) | 123 (43.8) | 93 (33.1) | 65 (23.1) |
| By channel type |  |  |  |  |  |  |  |
| All traditional media | 1,151 (44.8) | 175 (15.3) | 498 (43.6) | 363 (31.8) | 361 (31.4) | 464 (40.3) | 326 (28.3) |
| All social media | 358 (13.9) | 76 (21.2) | 184 (51.4) | 98 (27.4) | 94 (35.7) | 112 (42.6) | 57 (21.7) |
| Both social and traditional media | 1,059 (41.2) | 170 (16.1) | 708 (66.9) | 181 (17.1) | 441 (41.6) | 425 (40.1) | 193 (18.2) |

**Table 2.**
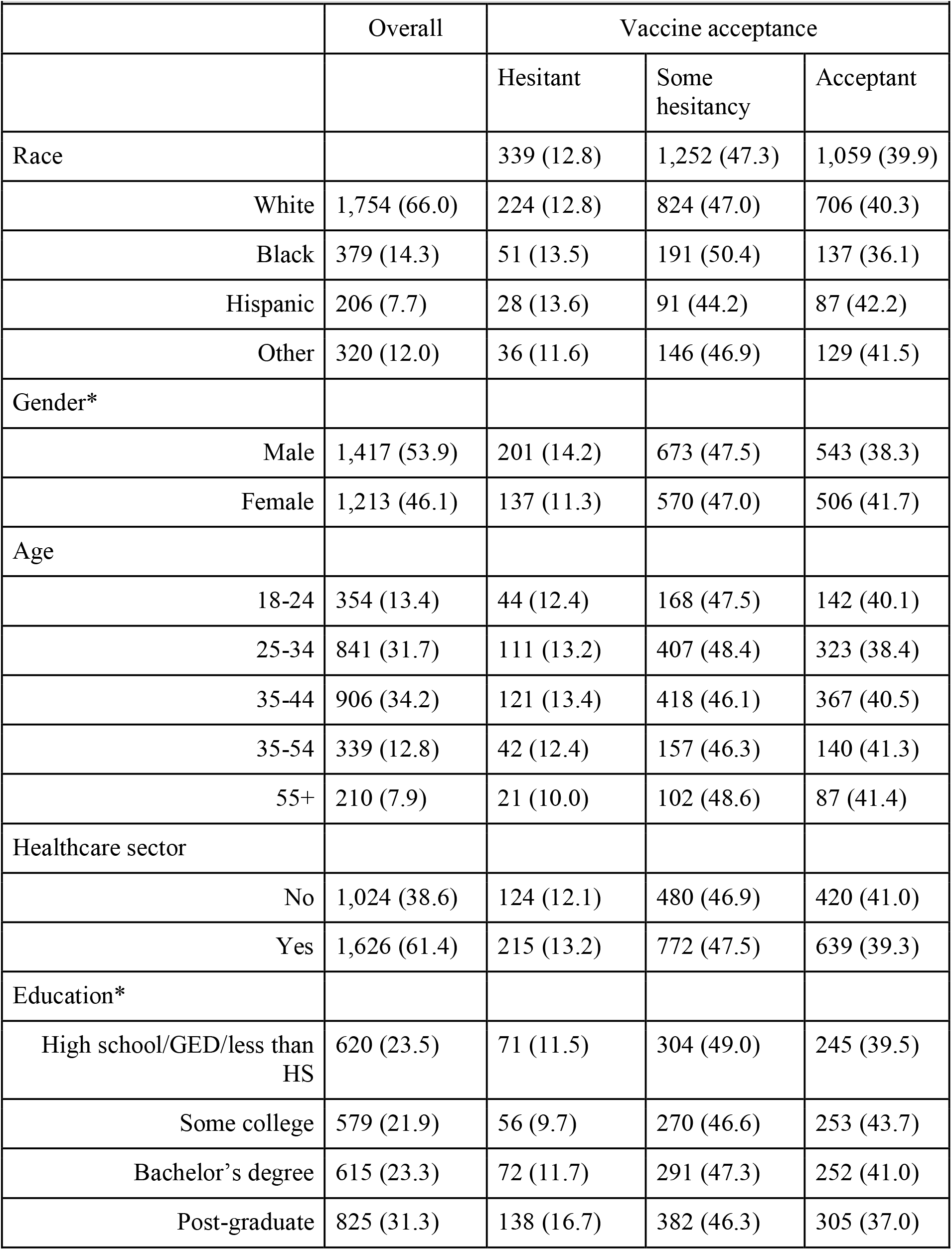
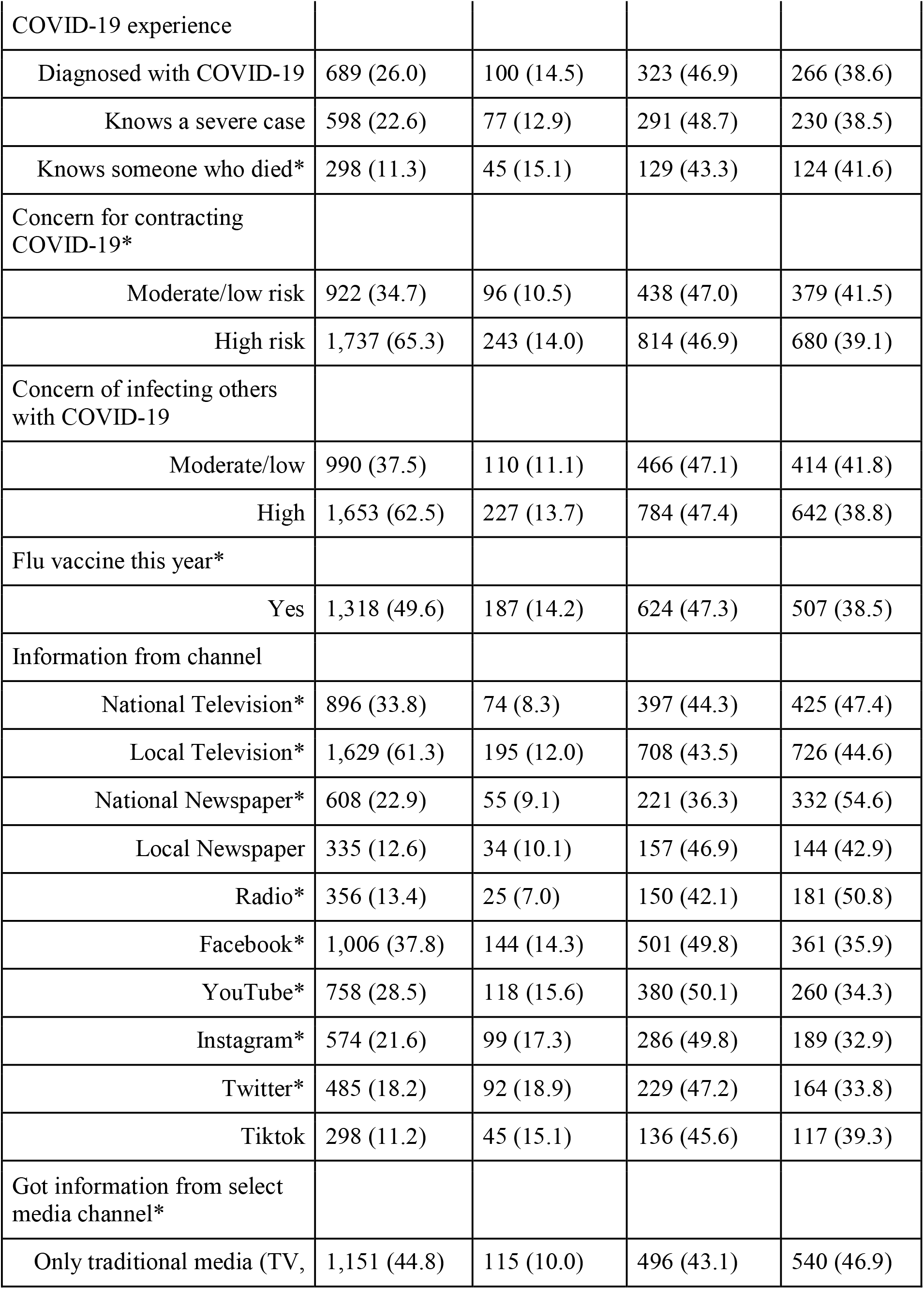
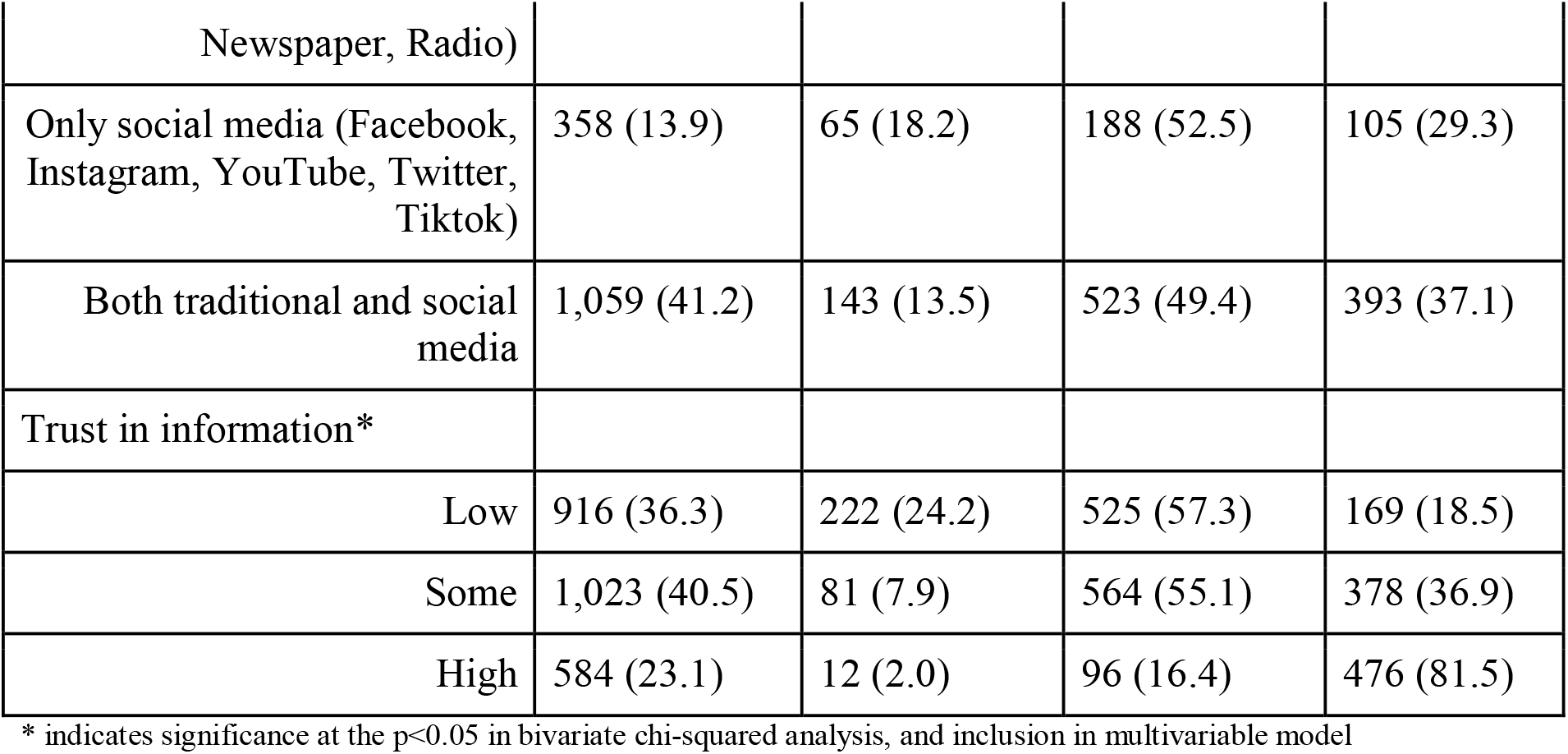
Univariate and bivariate analysis.

| **Table 2. Univariate and bivariate analysis** |  |  |  |  |
| --- | --- | --- | --- | --- |
|  | Overall | Vaccine acceptance |  |  |
|  |  | Hesitant | Some hesitancy | Acceptant |
| Race |  | 339 (12.8) | 1,252 (47.3) | 1,059 (39.9) |
| White | 1,754 (66.0) | 224 (12.8) | 824 (47.0) | 706 (40.3) |
| Black | 379 (14.3) | 51 (13.5) | 191 (50.4) | 137 (36.1) |
| Hispanic | 206 (7.7) | 28 (13.6) | 91 (44.2) | 87 (42.2) |
| Other | 320 (12.0) | 36 (11.6) | 146 (46.9) | 129 (41.5) |
| Gender* |  |  |  |  |
| Male | 1,417 (53.9) | 201 (14.2) | 673 (47.5) | 543 (38.3) |
| Female | 1,213 (46.1) | 137 (11.3) | 570 (47.0) | 506 (41.7) |
| Age |  |  |  |  |
| 18-24 | 354 (13.4) | 44 (12.4) | 168 (47.5) | 142 (40.1) |
| 25-34 | 841 (31.7) | 111 (13.2) | 407 (48.4) | 323 (38.4) |
| 35-44 | 906 (34.2) | 121 (13.4) | 418 (46.1) | 367 (40.5) |
| 35-54 | 339 (12.8) | 42 (12.4) | 157 (46.3) | 140 (41.3) |
| 55+ | 210 (7.9) | 21 (10.0) | 102 (48.6) | 87 (41.4) |
| Healthcare sector |  |  |  |  |
| No | 1,024 (38.6) | 124 (12.1) | 480 (46.9) | 420 (41.0) |
| Yes | 1,626 (61.4) | 215 (13.2) | 772 (47.5) | 639 (39.3) |
| Education* |  |  |  |  |
| High school/GED/less than HS | 620 (23.5) | 71 (11.5) | 304 (49.0) | 245 (39.5) |
| Some college | 579 (21.9) | 56 (9.7) | 270 (46.6) | 253 (43.7) |
| Bachelor's degree | 615 (23.3) | 72 (11.7) | 291 (47.3) | 252 (41.0) |
| Post-graduate | 825 (31.3) | 138 (16.7) | 382 (46.3) | 305 (37.0) |
| COVID-19 experience |  |  |  |  |
| Diagnosed with COVID-19 | 689 (26.0) | 100 (14.5) | 323 (46.9) | 266 (38.6) |
| Knows a severe case | 598 (22.6) | 77 (12.9) | 291 (48.7) | 230 (38.5) |
| Knows someone who died* | 298 (11.3) | 45 (15.1) | 129 (43.3) | 124 (41.6) |
| Concern for contracting COVID-19* |  |  |  |  |
| Moderate/low risk | 922 (34.7) | 96 (10.5) | 438 (47.0) | 379 (41.5) |
| High risk | 1,737 (65.3) | 243 (14.0) | 814 (46.9) | 680 (39.1) |
| Concern of infecting others with COVID-19 |  |  |  |  |
| Moderate/low | 990 (37.5) | 110 (11.1) | 466 (47.1) | 414 (41.8) |
| High | 1,653 (62.5) | 227 (13.7) | 784 (47.4) | 642 (38.8) |
| Flu vaccine this year* |  |  |  |  |
| Yes | 1,318 (49.6) | 187 (14.2) | 624 (47.3) | 507 (38.5) |
| Information from channel |  |  |  |  |
| National Television* | 896 (33.8) | 74 (8.3) | 397 (44.3) | 425 (47.4) |
| Local Television* | 1,629 (61.3) | 195 (12.0) | 708 (43.5) | 726 (44.6) |
| National Newspaper* | 608 (22.9) | 55 (9.1) | 221 (36.3) | 332 (54.6) |
| Local Newspaper | 335 (12.6) | 34 (10.1) | 157 (46.9) | 144 (42.9) |
| Radio* | 356 (13.4) | 25 (7.0) | 150 (42.1) | 181 (50.8) |
| Facebook* | 1,006 (37.8) | 144 (14.3) | 501 (49.8) | 361 (35.9) |
| YouTube* | 758 (28.5) | 118 (15.6) | 380 (50.1) | 260 (34.3) |
| Instagram* | 574 (21.6) | 99 (17.3) | 286 (49.8) | 189 (32.9) |
| Twitter* | 485 (18.2) | 92 (18.9) | 229 (47.2) | 164 (33.8) |
| Tiktok | 298 (11.2) | 45 (15.1) | 136 (45.6) | 117 (39.3) |
| Got information from select media channel* |  |  |  |  |
| Only traditional media (TV, | 1,151 (44.8) | 115 (10.0) | 496 (43.1) | 540 (46.9) |
| Newspaper, Radio) |  |  |  |  |
| Only social media (Facebook, Instagram, YouTube, Twitter, Tiktok) | 358 (13.9) | 65 (18.2) | 188 (52.5) | 105 (29.3) |
| Both traditional and social media | 1,059 (41.2) | 143 (13.5) | 523 (49.4) | 393 (37.1) |
| Trust in information* |  |  |  |  |
| Low | 916 (36.3) | 222 (24.2) | 525 (57.3) | 169 (18.5) |
| Some | 1,023 (40.5) | 81 (7.9) | 564 (55.1) | 378 (36.9) |
| High | 584 (23.1) | 12 (2.0) | 96 (16.4) | 476 (81.5) |

\* indicates significance at the $p < 0.05$ in bivariate chi-squared analysis, and inclusion in multivariable model

### Descriptive statistics

Table 1 describes the proportion of users of each information channel, the variations in attitude towards the vaccine across channels, as well as the level of trust in information among channels. The majority of respondents reported getting COVID-19 vaccine information from local TV (61%). The next highest source was Facebook (37.8%), followed by National TV, YouTube, National Newspaper, Instagram, Twitter, Radio, Local Newspaper, and Tiktok. Overall, 53.6% of respondents reported that the COVID-19 vaccine information they saw was mostly positive, 25.3% reported seeing it as neither positive or negative, 16.3% of respondents reported that what they heard about the was mostly negative, and 4.8% reported not seeing vaccine-related information on any channel (data not shown). The quality ratings of the information varied across channels. In contrast though, among those who used social media channels, the attitude toward the vaccine among those channels appeared to be relatively positive. The majority of respondents indicated that what they heard about the vaccine was positive, compared to negative or neutral (64-74%). Of those who used Twitter, nearly 75% reported the information they saw was positive (the highest among any channel), compared to just 47% on National TV and 48% in National Newspapers. Overall trust in information was high for 23% of respondents, moderate for 40.5%, and low for 36.3%. Though the majority reported overall what they heard was positive information, those who reported the information they saw was generally negative on the vaccine had higher levels of trust in information (20% had high trust in positive information vs. 31% had high trust in negative information, data not shown in table). The level of trust in vaccine information was highest for National newspapers and radio (approximately 34%).

Also, in Table 1, there were significant differences (p < 0.001) in both attitude of information and level of trust by channel type. Those who used only traditional media reported seeing information that was neither positive or negative at much higher rates (31.2%) than those that used social media or traditional media, and those that used both types of channels reported information was mostly positive (66.9%). Echoing the theme as looking at the individual channels, trust was higher in information from traditional media channels compared to social media channels, or a mix of the two sources. Figure 1 illustrates the level of trust in COVID-19 vaccine information across the channels described.

**Figure 1.**
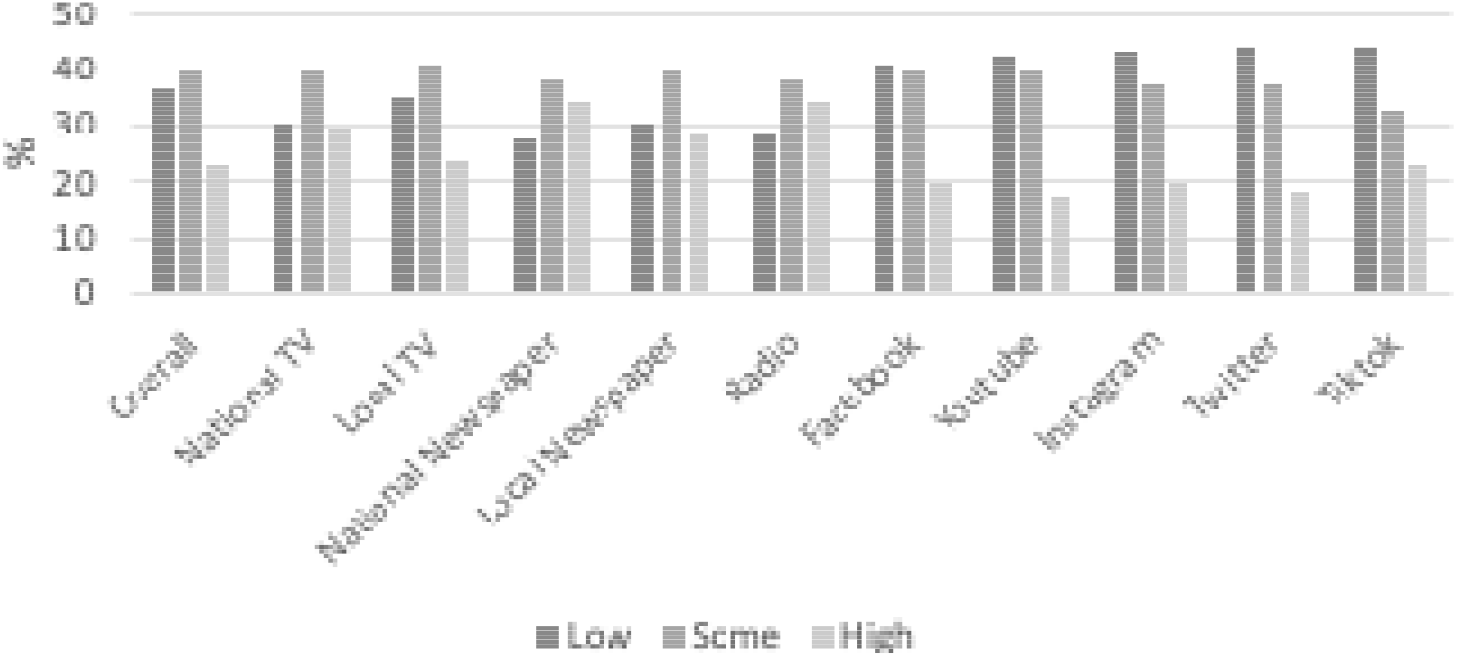
Trust in COVID-19 Information by Information Channel.

### Bivariate and Multivariable Regression Analysis

Table 2 presents the bivariate analysis among variables and vaccine acceptance. There were statistically significant differences in vaccine acceptance by gender, education, knowing someone who had died of COVID-19, level of perceived risk for contracting COVID-19, having received the flu vaccine, getting information from national TV, local TV, national newspaper, radio, Facebook, YouTube, Instagram, and Twitter, as well as level of trust in information. There was no significant difference in vaccination acceptance based on prior COVID-19 diagnosis. Females were more likely to be very likely to get the vaccine compared to males (41.7% vs. 38.0%), while those with post-graduate education were least likely to get the vaccine (37.0%) compared to other levels of education (Bachelor’s degree, 41.0%, Some-college 43.7%, and High school education or less 39.5%). A higher proportion of those who knew someone who had died of COVID-19 were very likely to get the vaccine. Those with higher perceived risk of contracting COVID-19, as well as those who had received the flu vaccine, were more hesitant. As shown in Table 2, a higher proportion of individuals who had gotten information from a national TV (47.4%), local TV (44.6%), national newspaper (54.6%), and radio (50.8) were vaccine acceptant compared to those who had not. Aggregating the channels of interest, there was a statistically significant difference in vaccine acceptance among those who had exclusively gotten information only from traditional media (46.9%), only from social media (29.3%), or both types of channels (37.1%). Those with high trust in COVID-19 vaccine information were more likely to get vaccinated (81.5%).

Table 3 shows the results of the first multinomial multivariable logistic regression analyzing the specific types of channels on vaccine acceptance. The generalized Hosmer– Lemeshow goodness-of-fit test for multinomial logistic regression models confirmed the model was a good fit for the data (p=0.787). Three channels of information were significant predictors of being very likely to take the vaccine: those that got information from national TV were 25% more vaccine acceptant (Relative Risk Ratio (RRR 1.25, CI 1.02-1.53)), those that got information from local TV were 75% more likely (RRR 1.75, CI 1.40-2.19), and those that got information from a national newspaper were over 80% more acceptant (RRR 1.81, CI 1.45-2.24) to do so, compared to those who did not use the source. Trust in information greatly increased the relative risk of vaccine acceptance. Some trust in COVID-19 vaccine information doubled the relative risk of vaccine acceptance (RRR 2.01, CI 1.61-2.51) while high trust in information was fifteen times more likely (RRR 15.04, CI 11.26-20.09). Having a close family member or friend who had died of COVID-19, respondents were 47% more acceptant of the vaccine (OR 1.47, CI 1.08, 1.99). No other covariate increased the relative risk of vaccine acceptance. As shown in Table 4, we explored a second multivariable model to move beyond individual channels, to investigate whether there was an effect due to using a particular type of media channel (traditional vs. social vs. both). While there were no statistically significant changes to the relative risk of vaccine acceptance due to any other variable, there was a clear decrease in acceptance if individuals exclusively used social media (RRR 0.45, CI 0.32-0.64) or used both social media and traditional media channels (RRR 0.81, CI 0.66-1.00) compared to those who only used traditional media.

**TABLE 3.**
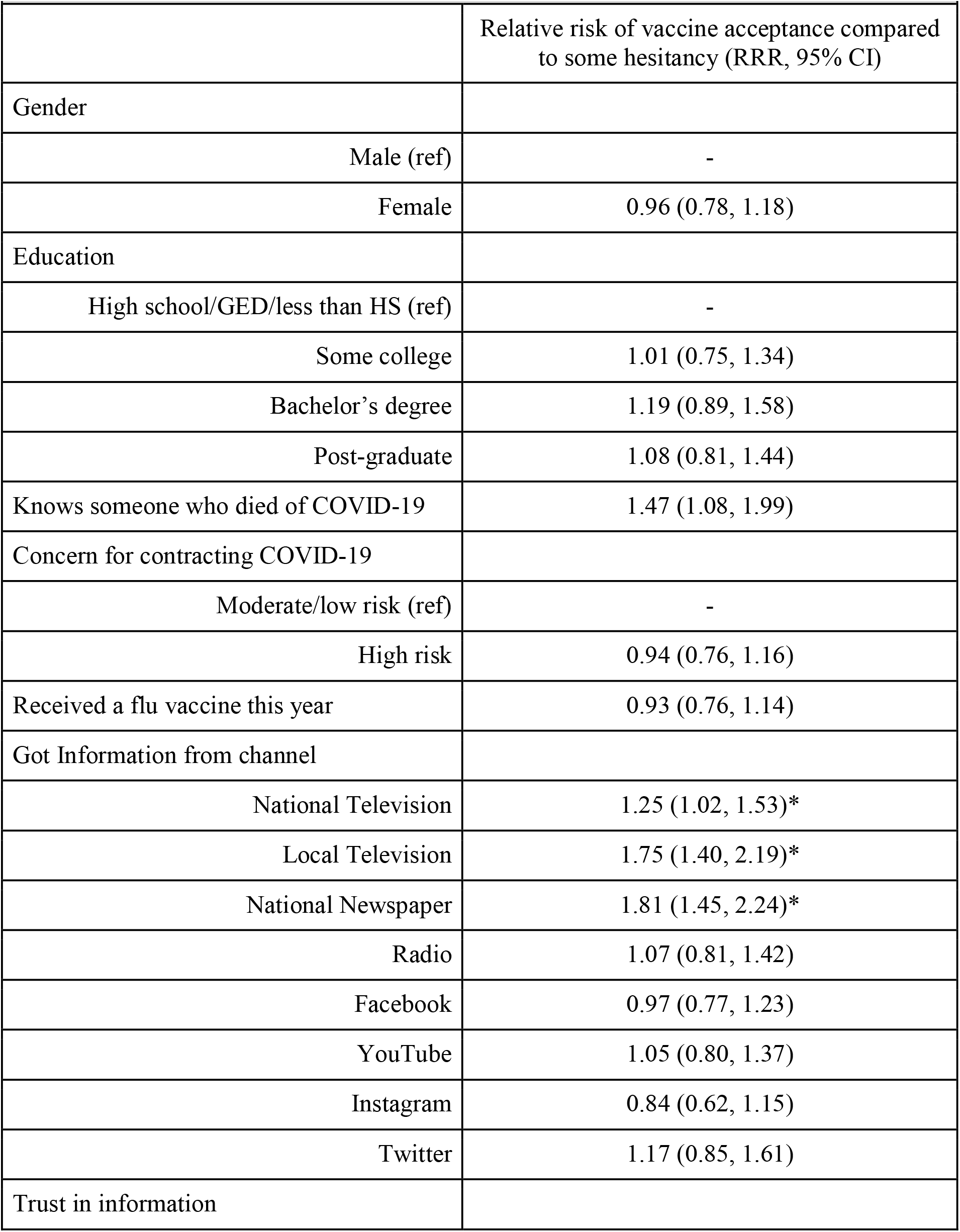
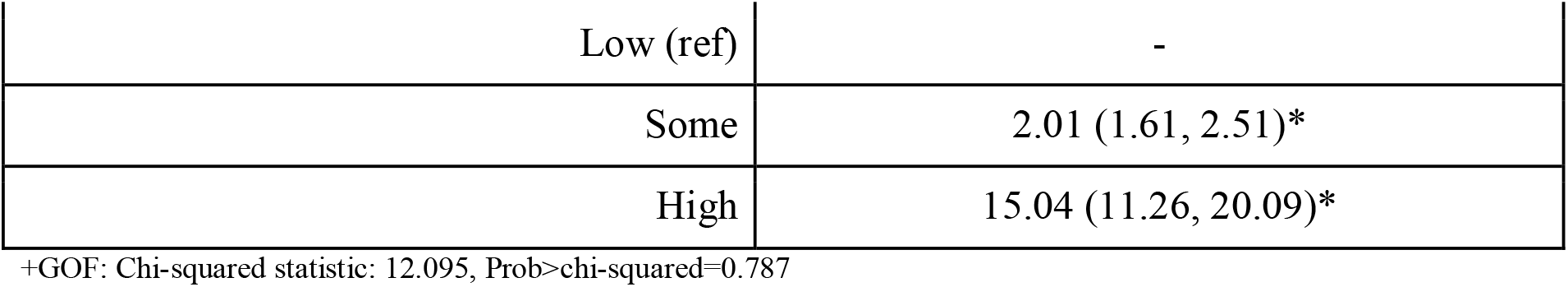
Multivariable multinomial regression of vaccine acceptance.

| **TABLE 3. Multivariable multinomial regression of vaccine acceptance** |  |
| --- | --- |
|  | Relative risk of vaccine acceptance compared to some hesitancy (RRR, 95% CI) |
| Gender |  |
| Male (ref) | - |
| Female | 0.96 (0.78, 1.18) |
| Education |  |
| High school/GED/less than HS (ref) | - |
| Some college | 1.01 (0.75, 1.34) |
| Bachelor's degree | 1.19 (0.89, 1.58) |
| Post-graduate | 1.08 (0.81, 1.44) |
| Knows someone who died of COVID-19 | 1.47 (1.08, 1.99) |
| Concern for contracting COVID-19 |  |
| Moderate/low risk (ref) | - |
| High risk | 0.94 (0.76, 1.16) |
| Received a flu vaccine this year | 0.93 (0.76, 1.14) |
| Got Information from channel |  |
| National Television | 1.25 (1.02, 1.53)* |
| Local Television | 1.75 (1.40, 2.19)* |
| National Newspaper | 1.81 (1.45, 2.24)* |
| Radio | 1.07 (0.81, 1.42) |
| Facebook | 0.97 (0.77, 1.23) |
| YouTube | 1.05 (0.80, 1.37) |
| Instagram | 0.84 (0.62, 1.15) |
| Twitter | 1.17 (0.85, 1.61) |
| Trust in information |  |
| Low (ref) | - |
| Some | 2.01 (1.61, 2.51)* |
| High | 15.04 (11.26, 20.09)* |

+GOF: Chi-squared statistic: 12.095, Prob>chi-squared=0.787

**TABLE 4.** Multivariable multinomial regression of likelihood of vaccine acceptance.

+GOF: Chi-squared statistic: 12.095, Prob>chi-squared=0.787
|  |  |
| --- | --- |
|  | Relative risk of vaccine acceptance compared to some hesitancy (RR, CI) |
| Gender |  |
| Male (ref) | - |
| Female | 0.97 (0.80, 1.19) |
| Education |  |
| High school/GED/less than HS (ref) | - |
| Some college | 1.06 (0.79, 1.40) |
| Bachelor's degree | 1.25 (0.94, 1.66) |
| Post-graduate | 1.11 (0.84, 1.47) |
| Knows someone who died of COVID-19 | 1.46 (1.07, 1.98) |
| Concern for contracting COVID-19 |  |
| Moderate/low risk (ref) | - |
| High risk | 0.98 (0.79, 1.21) |
| Received a flu vaccine this year | 0.94 (0.77, 1.14) |
| Got information from select media channel |  |
| Only traditional media (TV, Newspaper, Radio) (ref) | - |
| Only social media (Facebook, Instagram, YouTube, Twitter, Tiktok) | 0.45 (0.32, 0.64) |
| Both traditional and social media | 0.81 (0.66, 1.00) |
| Trust in information |  |
| Low (ref) | - |
| Some | 2.05 (1.65, 2.56) |
| High | 15.60 (11.69, 20.81) |

\*GOF: Chi-squared statistic: 8.663, Prob>chi-squared=0.927

## Discussion

How to increase vaccine uptake in a population that has relatively high levels of vaccine hesitancy is an ongoing public health challenge. Our analysis compares being “very likely to take the vaccine,” which we defined as “acceptant” to “somewhat likely or unlikely”, which we defined as “hesitant” enabling us to isolate the factors that increase the relative risk of vaccine uptake relative to being hesitant. Our sample had high levels of exposure to COVID-19 (26% reported having had the virus) compared to the average US population estimates of 7% with ranges estimated between 1 and 23%, likely due to workforce exposure (27). Overall, respondents were also more likely to be willing to get the vaccine compared to other poll results from the same time period (1).

The results show that the majority of respondents are using traditional media to obtain information on the COVID-19 vaccine in some way (86%) and use of traditional media sources was found to increase the likelihood of vaccination. In particular, obtaining information from TV, both local and national, and national newspapers, increased acceptance of vaccination (regardless of how much the channel is trusted). These channels are likely adhering to high quality sources, sharing fact-based vaccine information linked to governmental, healthcare, or academic data and reports, and are clearly crucial channels to promote immunization programs (28). Traditional media channels should continue to promote vaccination information to their viewers and readers.

In contrast, there did not seem to be independent effects of receiving information from a given social media platform. However, the results also show that those who are less likely to get the vaccine are using social media as their sole source of information, or as at least one of their sources of information. Social media platforms therefore have an opportunity to consider how information on those platforms can be crafted to encourage vaccination. Given the vulnerability of social media channels to exploitation by bad actors, and their diminished trustworthiness relative to legacy media, it may be necessary to draw on traditional channels to pursue persuasive campaigns more pointedly addressing the needs of audiences with neutral and oppositional attitudes toward the COVID-19 vaccine. The same customization and targeted marketing capabilities that enable exploitation by bad actors on social media might also offer opportunities to address these needs using a one-step communicative approach whereby information is targeted to an individual across multiple platforms (16, 17).

We also considered if trust was a potential mediator of the relationship between channel and hesitancy, hypothesizing if a channel was trusted, the relationship to vaccination likelihood would change. To identify if trust in a particular channel was driving the effect of that channel, we tested interaction terms for each traditional channel that was significant, as well as by channel types, but found no significant interaction effects between type of media channel and trust. This suggests it is not just trust in a given channel that is driving its impact on vaccination likelihood. We hypothesize it is the combination of familiarity in traditional media and perceived credibility of the sources they cite such as public health experts or academic leaders that is contributing to the effect of these channels. Traditional media channels are now accessible online, and often they have made their content on the COVID-19 vaccine free for public viewing.

### Limitations

This is a cross-sectional study and cannot disentangle if those that are already more likely to take the vaccine are drawn to traditional media channels, while those that are more hesitant are using social media or other channels we did not include in our analysis. Further longitudinal analysis is necessary to explore this issue. Additionally, TV and newspaper channels may be promoted on social media channels such as when an individual sees a given newspaper’s story being shared on Facebook. This analysis cannot separate out the impact of this exchange.

Respondents were also not asked exclusively to select a particular channel of information. This paper also did not evaluate the cumulative effect of getting information from multiple channels or the frequency of information from a given channel. Additionally, this is not a representative sample, but rather a convenience sample of workers in potential vaccine priority groups in the US. The results may not be generalizable beyond this sample.

## Conclusions

The COVID-19 vaccine rollout is already underway. Anecdotal reports suggest some of those who are eligible for the vaccine are refusing to get vaccinated (29, 30). This paper demonstrates it is useful to draw on traditional media sources to discuss the COVID-19 vaccine. We also show that because vaccine-hesitant individuals are more likely to identify social media as their sole source of information, social media platforms have a particularly important role to play in addressing vaccine hesitancy.

## Data Availability

Data are available upon request from the authors.

## Notes

### Competing Interest Statement

The authors have declared no competing interest.

### Funding Statement

Research reported in this publication was supported by Jigsaw, Google. The content is solely the responsibility of the authors and does not necessarily represent the official views of the sponsor.

### Author Declarations

The study was approved by the Institutional Review Board at the Harvard T.H. Chan School of Public Health Office of Regulatory Affairs and Research Compliance 90 Smith Street, 3rd Floor Boston, MA 02120 Federal wide Assurance # FWA00002642

